# Myocardial infarction with non-obstructive coronary arteries: a single-center retrospective study by sex and race

**DOI:** 10.1101/2023.06.08.23291173

**Authors:** Ghenekaro Esin, Christine Hsueh, Thomas Breen, Mauro Gitto, Miriam Katz, Martha Gulati, Quinn Capers, Harmony R. Reynolds, Annabelle S. Volgman, S. Elissa Altin

**Author notes:** as co first (Ghenekaro Esin, Christine Hsueh). Address for Correspondence: S. Elissa Altin, MD Section of Cardiovascular Medicine Department of Internal Medicine Yale School of Medicine 789 Howard Ave New Haven, CT 06519.

## Abstract

**Background:** In myocardial infarction with non-obstructive coronary arteries (MINOCA), there are limited patient-level data on outcomes by sex and race.

**Objective:** Assess baseline demographics and 3-year outcomes by sex and race for MINOCA patients.

**Methods:** Patients admitted to a single center with acute myocardial infarction (AMI) between January 1, 2012 and December 31, 2018, were identified by chart and angiographic review. The primary outcome was nonfatal MI with secondary outcomes including non-fatal cerebrovascular accident (CVA), chest pain readmission, and repeat coronary angiography.

**Results:** During the study period, 304 patients were admitted with MINOCA. The cohort was predominantly female (66.4%), and women were significantly older (64.6 vs. 59.2). One-sixth of the total population were Black patients, and nearly half of Black patients (47.2%) were male. Prior CVA (19.7%) and comorbid anxiety, depression, or post-traumatic stress disorder (41.1%) were common. Rates of non-fatal myocardial infarction (MI) were 6.3% without difference by sex or race. For secondary outcomes, rates of CVA were 1.7%, chest pain readmission were 22.4%, and repeat angiography were 8.9%. Men were significantly more likely to have repeat angiography (13.7% vs. 6.4%), and Black patients more likely to be readmitted for angina (34.0% vs. 19.1%). Over one-quarter of patients underwent repeat stress testing, with 8.9% ultimately undergoing repeat angiograms and low numbers (0.7%) undergoing revascularization. Men were more likely to be referred for a repeat angiogram (13.7% vs. 6.4%, p=0.035). In multivariate analysis, Black race (OR 2.31 [95% CI (1.06-5.03)] was associated with an increased risk of readmission for angina, while female sex was associated with decreased odds of repeat angiography (OR 0.36 [95% CI (0.14-0.90)] and current smoking was associated with increased odds of repeat angiography (OR 4.07 [95% CI (1.02-16.29)] along with hyperlipidemia (OR 4.65 [95% CI (1.22-17.7)].

**Conclusion:** White women presented more frequently with MINOCA than White men, however Black men are equally as affected as Black women. Rates of non-fatal MI were low without statistical difference by sex or race.

## Introduction

Myocardial infarction with non-obstructive coronary artery disease (MINOCA) represents a clinical presentation of acute myocardial infarction (AMI) where the burden of epicardial coronary artery disease (CAD) insufficiently explains the extent of ischemia. MINOCA is estimated to occur in approximately 5% of all AMI presentations in the United States^1, 2^ and represents heterogeneous underlying pathophysiologic processes. Prior studies have identified clinical presentation, characteristics, and outcomes of patients with MINOCA, mainly focusing on an at-risk population of younger patients and women with a high cardiac risk profile despite no flow-limiting epicardial CAD. However, there are limited data regarding demographic differences and rates of major adverse cardiovascular events (MACE) by sex and race in a contemporary cohort. Prior large retrospective registry studies report a 5% MACE rate without any difference in in-hospital mortality by sex^1, 3^.

The aim of this retrospective study was to assess baseline demographic and clinical presentations by sex and race as well as long-term outcome rates in a contemporary cohort of patients presenting with MINOCA across a large health system.

## Methods

### Study Population

We conducted a retrospective study of patients diagnosed with MINOCA at Yale-New Haven Hospital between January 1, 2012 and December 31, 2018. We reviewed 1,124 patients who presented with AMI and did not receive percutaneous coronary intervention or coronary artery bypass surgery (see **Supplemental Table 1** for codes) and then identified living patients with MINOCA by chart review (**Figure 1**). The diagnostic criteria for MINOCA were defined according to the standardized definition in patients with AMI from the “Fourth Universal Definition of Myocardial Infarction” and clinical evidence of one of the following: symptoms of myocardial ischemia, new ischemic electrocardiographic changes, development of pathological Q waves, imaging evidence of ischemia with no obstructive disease on angiography with diameter stenosis ≤50% angiographic stenosis, and no other specific diagnosis to explain the clinical presentation^2^. Notably, we excluded patients with sepsis, pulmonary embolism, myocarditis, type II non-ST elevation myocardial infarction, chronic total occlusion, prior coronary stents, concurrent cocaine use or cocaine-induced vasospasm, Takotsubo cardiomyopathy, and spontaneous coronary artery dissection (SCAD).

**Figure 1.**
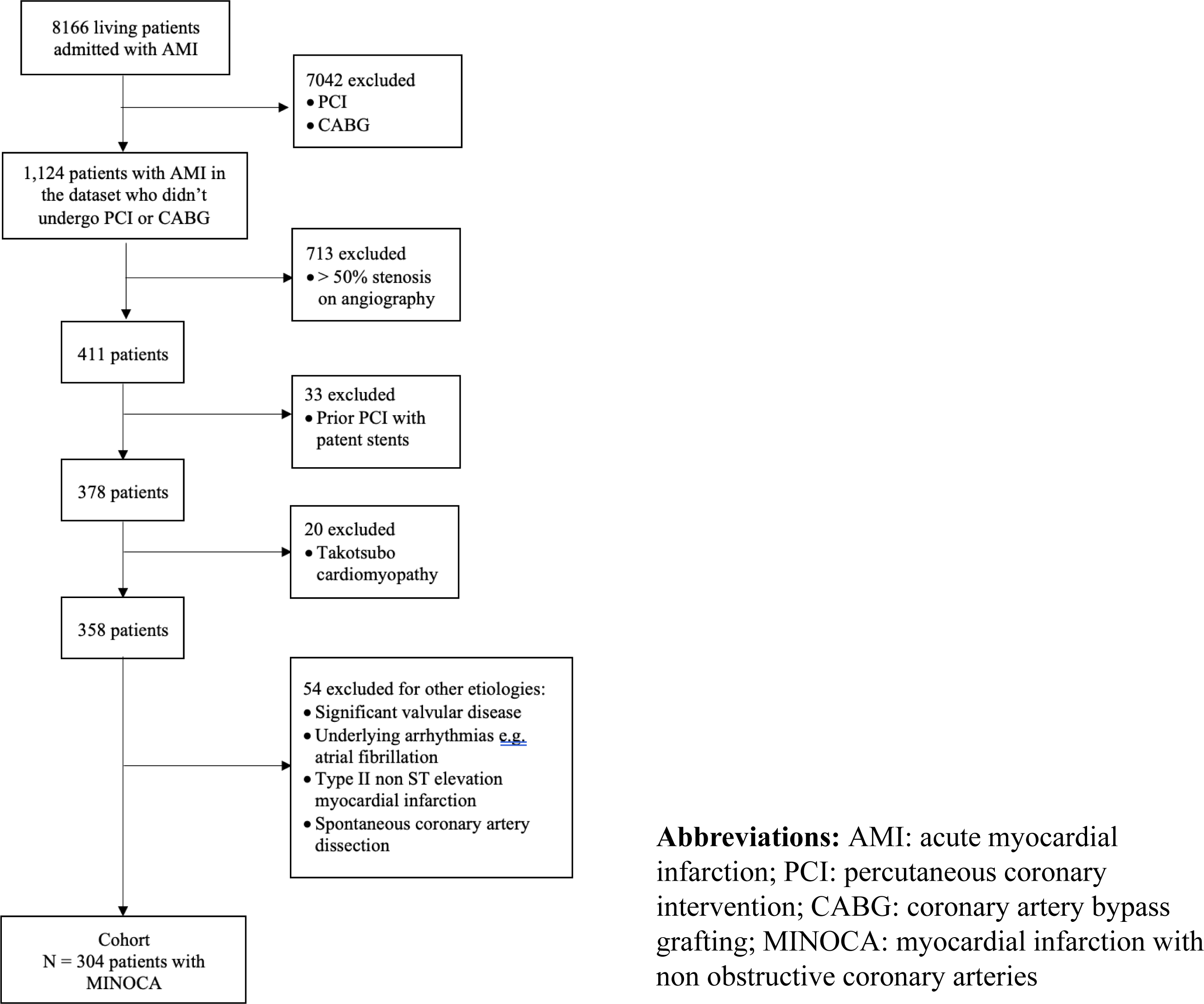
Flowsheet showing selection criteria for the final study cohort.

Clinical and historical data were abstracted by medical chart review. Information on cardiac risk factors, admission diagnoses, discharge medications, and follow-up was also collected. Based on chart review, the self-identified race was categorized as Black, White, Asian or other/unknown, and ethnicity as Hispanic or non-Hispanic. The IRB committee approved this study at Yale-New Haven Hospital.

### Outcomes

The primary outcome was nonfatal MI. Secondary outcomes were non-fatal cerebrovascular accident (CVA), chest pain readmission, repeat coronary angiography, referral to cardiology follow-up, and left ventricular ejection fraction (LVEF) at follow-up.

### Statistics

Data are reported as mean ± standard deviation for continuous variables and number (%) for categorical variables. Comparisons were made using the student’s T-test for continuous variables and Pearson’s chi-squared test for categorical variables. A multivariate logistic regression model that included cardiovascular risk factors plausibly associated with cardiovascular outcomes was developed to identify independent predictors of primary and secondary outcomes. Statistical analyses were performed using JMP 16 (Cary, NC).

## Results

### Baseline Demographics by Sex and Race

During the study period, 8,166 patients were admitted with AMI, and of these 304 (3.7%) met criteria for MINOCA. In this cohort of 304 patients with MINOCA, there was a 2:1 predominance of women over men (66.4% women vs. 33.6% men, **Table 1**). Of the MINOCA cases, 17.4% occurred in Black patients, with MINOCA occurring equally in Black men and Black women (52.8 % vs 47.2%, p=0.58). Women with MINOCA were older than men (64.6 ± 13.2 vs. 59.2 ± 13.6, p=0.001), with no significant differences in rates of cardiac risk factors or other comorbidities. Black patients with MINOCA were more likely to have higher body mass index than Whites (31.6 ± 8.0 vs. 28.6 ± 8.2, p=0.02) and hypertension (37.7% vs. 23%, p=0.03). There were no other significant differences in cardiovascular risk factors by sex or race (**Table 1**).

**Table 1:**
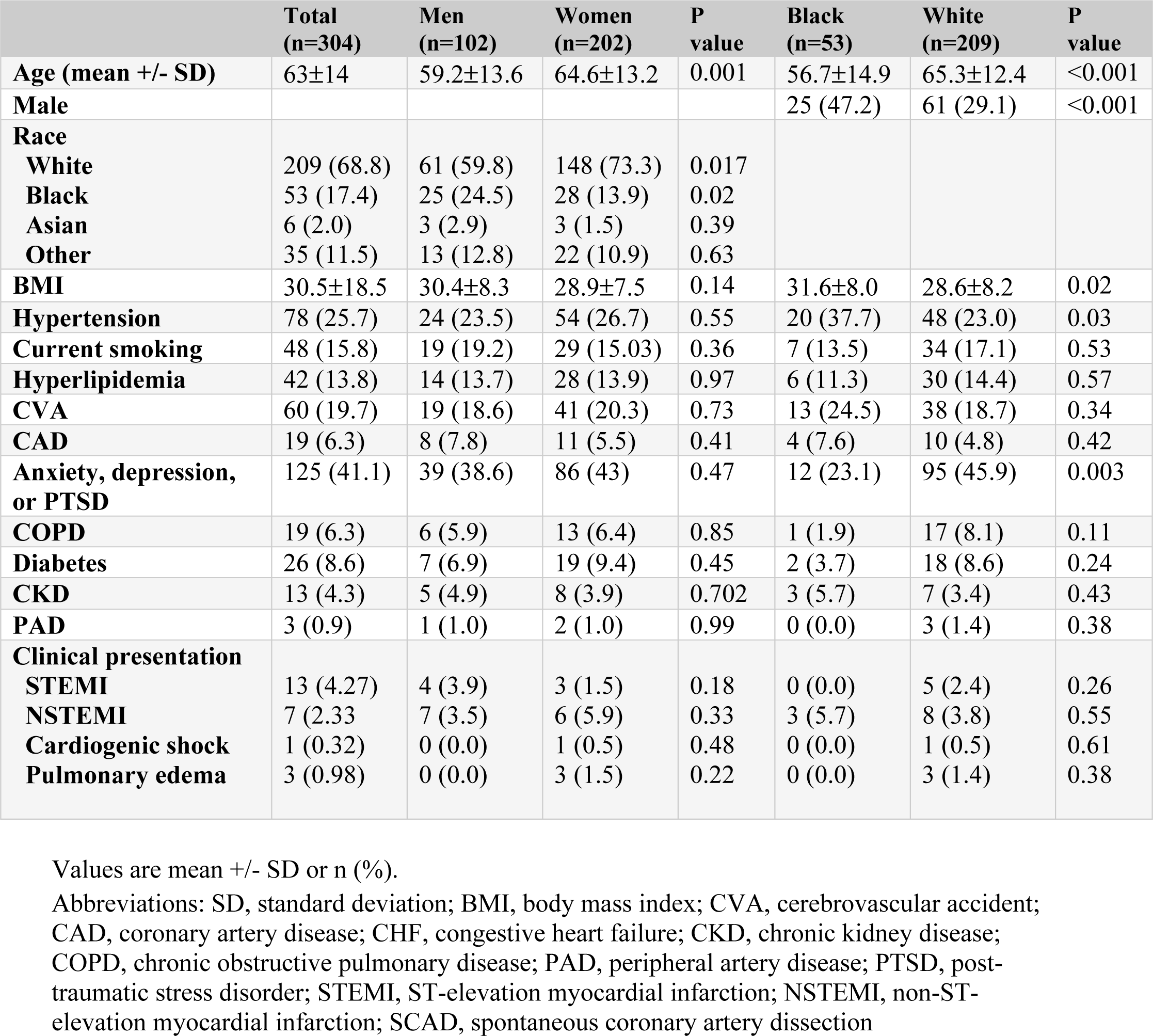
Baseline demographics and risk factors by sex and race

|  | **Total**<br>**(n=304)** | **Men**<br>**(n=102)** | **Women**<br>**(n=202)** | **P**<br>**value** | **Black**<br>**(n=53)** | **White**<br>**(n=209)** | **P**<br>**value** |
| --- | --- | --- | --- | --- | --- | --- | --- |
| **Age (mean +/- SD)** | 63±14 | 59.2±13.6 | 64.6±13.2 | 0.001 | 56.7±14.9 | 65.3±12.4 | <0.001 |
| **Male** |  |  |  |  | 25 (47.2) | 61 (29.1) | <0.001 |
| **Race** |  |  |  |  |  |  |  |
| **White** | 209 (68.8) | 61 (59.8) | 148 (73.3) | 0.017 |  |  |  |
| **Black** | 53 (17.4) | 25 (24.5) | 28 (13.9) | 0.02 |  |  |  |
| **Asian** | 6 (2.0) | 3 (2.9) | 3 (1.5) | 0.39 |  |  |  |
| **Other** | 35 (11.5) | 13 (12.8) | 22 (10.9) | 0.63 |  |  |  |
| **BMI** | 30.5±18.5 | 30.4±8.3 | 28.9±7.5 | 0.14 | 31.6±8.0 | 28.6±8.2 | 0.02 |
| **Hypertension** | 78 (25.7) | 24 (23.5) | 54 (26.7) | 0.55 | 20 (37.7) | 48 (23.0) | 0.03 |
| **Current smoking** | 48 (15.8) | 19 (19.2) | 29 (15.03) | 0.36 | 7 (13.5) | 34 (17.1) | 0.53 |
| **Hyperlipidemia** | 42 (13.8) | 14 (13.7) | 28 (13.9) | 0.97 | 6 (11.3) | 30 (14.4) | 0.57 |
| **CVA** | 60 (19.7) | 19 (18.6) | 41 (20.3) | 0.73 | 13 (24.5) | 38 (18.7) | 0.34 |
| **CAD** | 19 (6.3) | 8 (7.8) | 11 (5.5) | 0.41 | 4 (7.6) | 10 (4.8) | 0.42 |
| **Anxiety, depression,**<br>**or PTSD** | 125 (41.1) | 39 (38.6) | 86 (43) | 0.47 | 12 (23.1) | 95 (45.9) | 0.003 |
| **COPD** | 19 (6.3) | 6 (5.9) | 13 (6.4) | 0.85 | 1 (1.9) | 17 (8.1) | 0.11 |
| **Diabetes** | 26 (8.6) | 7 (6.9) | 19 (9.4) | 0.45 | 2 (3.7) | 18 (8.6) | 0.24 |
| **CKD** | 13 (4.3) | 5 (4.9) | 8 (3.9) | 0.702 | 3 (5.7) | 7 (3.4) | 0.43 |
| **PAD** | 3 (0.9) | 1 (1.0) | 2 (1.0) | 0.99 | 0 (0.0) | 3 (1.4) | 0.38 |
| **Clinical presentation** |  |  |  |  |  |  |  |
| **STEMI** | 13 (4.27) | 4 (3.9) | 3 (1.5) | 0.18 | 0 (0.0) | 5 (2.4) | 0.26 |
| **NSTEMI** | 7 (2.33) | 7 (3.5) | 6 (5.9) | 0.33 | 3 (5.7) | 8 (3.8) | 0.55 |
| **Cardiogenic shock** | 1 (0.32) | 0 (0.0) | 1 (0.5) | 0.48 | 0 (0.0) | 1 (0.5) | 0.61 |
| **Pulmonary edema** | 3 (0.98) | 0 (0.0) | 3 (1.5) | 0.22 | 0 (0.0) | 3 (1.4) | 0.38 |

Values are mean +/- SD or n (%).
Abbreviations: SD, standard deviation; BMI, body mass index; CVA, cerebrovascular accident; CAD, coronary artery disease; CHF, congestive heart failure; CKD, chronic kidney disease; COPD, chronic obstructive pulmonary disease; PAD, peripheral artery disease; PTSD, post-traumatic stress disorder; STEMI, ST-elevation myocardial infarction; NSTEMI, non-ST-elevation myocardial infarction; SCAD, spontaneous coronary artery dissection

In this cohort, 25.7% had hypertension, 13.8% had hyperlipidemia, 4.3% had chronic kidney disease (CKD), 0.9% had peripheral arterial disease (PAD), and 8.6% had diabetes mellitus, with no difference based on sex. Black patients with MINOCA had significantly more hypertension and obesity. Over 40% of patients had a diagnosis of anxiety, depression, or post-traumatic stress disorder (PTSD) at the time of presentation with MINOCA without difference by sex. However, Black patients with MINOCA were significantly less likely to have a diagnosis of anxiety, depression, or PTSD (23.1% vs. 45.9%, p=0.003) than White patients.

### In-hospital Complications and Evaluation

In-hospital complications for admission of AMI were rare (**Table 2**). There were no in-hospital deaths, 1 (0.3%) received a transfusion, and 5 (1.7%) patients suffered acute renal failure requiring dialysis. Three patients suffered an in-hospital diagnosis of CVA (1.0%). There were no significant differences based on sex or race. Referral for cardiac MRI was low overall at 6.3%, either during the index hospitalization or with readmission.

**Table 2:**
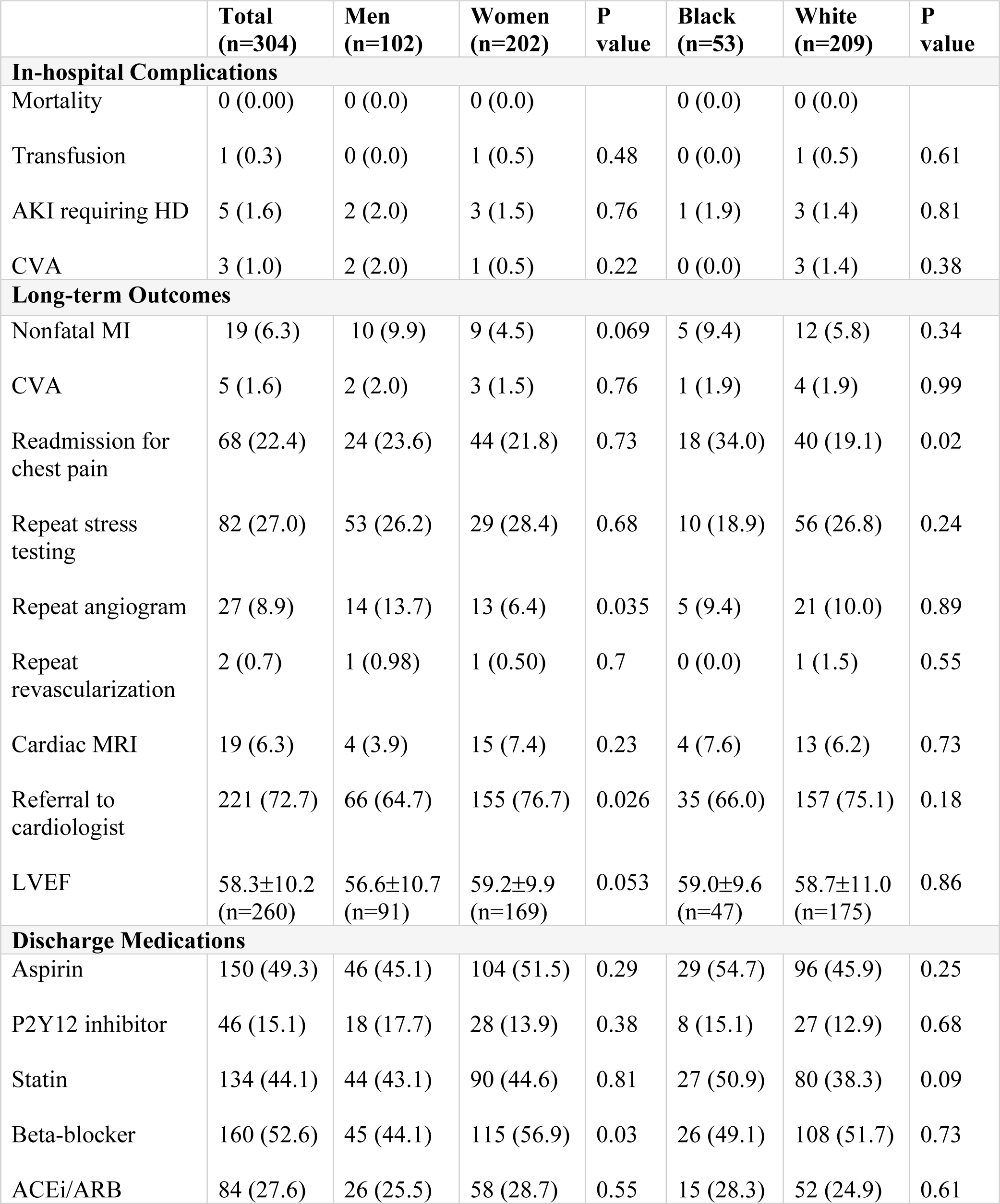
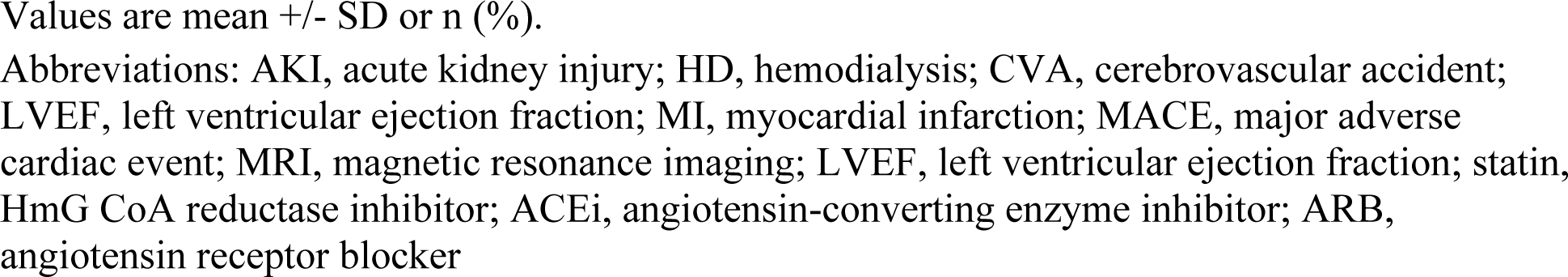
Outcomes and discharge medications stratified by sex and race

|  | **Total**<br>**(n=304)** | **Men**<br>**(n=102)** | **Women**<br>**(n=202)** | **P**<br>**value** | **Black**<br>**(n=53)** | **White**<br>**(n=209)** | **P**<br>**value** |
| --- | --- | --- | --- | --- | --- | --- | --- |
| **In-hospital Complications** |  |  |  |  |  |  |  |
| Mortality | 0 (0.00) | 0 (0.0) | 0 (0.0) |  | 0 (0.0) | 0 (0.0) |  |
| Transfusion | 1 (0.3) | 0 (0.0) | 1 (0.5) | 0.48 | 0 (0.0) | 1 (0.5) | 0.61 |
| AKI requiring HD | 5 (1.6) | 2 (2.0) | 3 (1.5) | 0.76 | 1 (1.9) | 3 (1.4) | 0.81 |
| CVA | 3 (1.0) | 2 (2.0) | 1 (0.5) | 0.22 | 0 (0.0) | 3 (1.4) | 0.38 |
| **Long-term Outcomes** |  |  |  |  |  |  |  |
| Nonfatal MI | 19 (6.3) | 10 (9.9) | 9 (4.5) | 0.069 | 5 (9.4) | 12 (5.8) | 0.34 |
| CVA | 5 (1.6) | 2 (2.0) | 3 (1.5) | 0.76 | 1 (1.9) | 4 (1.9) | 0.99 |
| Readmission for chest pain | 68 (22.4) | 24 (23.6) | 44 (21.8) | 0.73 | 18 (34.0) | 40 (19.1) | 0.02 |
| Repeat stress testing | 82 (27.0) | 53 (26.2) | 29 (28.4) | 0.68 | 10 (18.9) | 56 (26.8) | 0.24 |
| Repeat angiogram | 27 (8.9) | 14 (13.7) | 13 (6.4) | 0.035 | 5 (9.4) | 21 (10.0) | 0.89 |
| Repeat revascularization | 2 (0.7) | 1 (0.98) | 1 (0.50) | 0.7 | 0 (0.0) | 1 (1.5) | 0.55 |
| Cardiac MRI | 19 (6.3) | 4 (3.9) | 15 (7.4) | 0.23 | 4 (7.6) | 13 (6.2) | 0.73 |
| Referral to cardiologist | 221 (72.7) | 66 (64.7) | 155 (76.7) | 0.026 | 35 (66.0) | 157 (75.1) | 0.18 |
| LVEF | 58.3±10.2<br>(n=260) | 56.6±10.7<br>(n=91) | 59.2±9.9<br>(n=169) | 0.053 | 59.0±9.6<br>(n=47) | 58.7±11.0<br>(n=175) | 0.86 |
| **Discharge Medications** |  |  |  |  |  |  |  |
| Aspirin | 150 (49.3) | 46 (45.1) | 104 (51.5) | 0.29 | 29 (54.7) | 96 (45.9) | 0.25 |
| P2Y12 inhibitor | 46 (15.1) | 18 (17.7) | 28 (13.9) | 0.38 | 8 (15.1) | 27 (12.9) | 0.68 |
| Statin | 134 (44.1) | 44 (43.1) | 90 (44.6) | 0.81 | 27 (50.9) | 80 (38.3) | 0.09 |
| Beta-blocker | 160 (52.6) | 45 (44.1) | 115 (56.9) | 0.03 | 26 (49.1) | 108 (51.7) | 0.73 |
| ACEi/ARB | 84 (27.6) | 26 (25.5) | 58 (28.7) | 0.55 | 15 (28.3) | 52 (24.9) | 0.61 |

Values are mean +/- SD or n (%).
Abbreviations: AKI, acute kidney injury; HD, hemodialysis; CVA, cerebrovascular accident; LVEF, left ventricular ejection fraction; MI, myocardial infarction; MACE, major adverse cardiac event; MRI, magnetic resonance imaging; LVEF, left ventricular ejection fraction; statin, HmG CoA reductase inhibitor; ACEi, angiotensin-converting enzyme inhibitor; ARB, angiotensin receptor blocker

### Long-term Outcomes

The primary outcome of nonfatal MI occurred in 19 (6.3%) of patients, without difference by sex or race, however there was a non-significant trend for men compared to women (9.9% vs. 4.5%, p=0.069) **(Table 1, Figure 2)**. For secondary outcomes, CVA occurred in 1.6% of patients without difference by sex or race. Over one-fifth of patients were readmitted for chest pain without difference by sex. However, Black patients were significantly more likely than White patients to be readmitted (34.0% vs. 19.1%, p=0.02) (**Figure 3**). Over one-quarter of patients underwent repeat stress testing, with 8.9% ultimately undergoing repeat angiograms and low numbers (0.7%) undergoing revascularization. Men were more likely to be referred for a repeat angiogram (13.7% vs. 6.4%, p=0.035). LVEF at follow-up was preserved with a mean of >55%. Referral rates to a cardiologist at discharge were high at 72.7%, with women more likely than men to be referred (76.7% vs. 64.7%, p=0.026), without difference by race.

**Figure 2.**
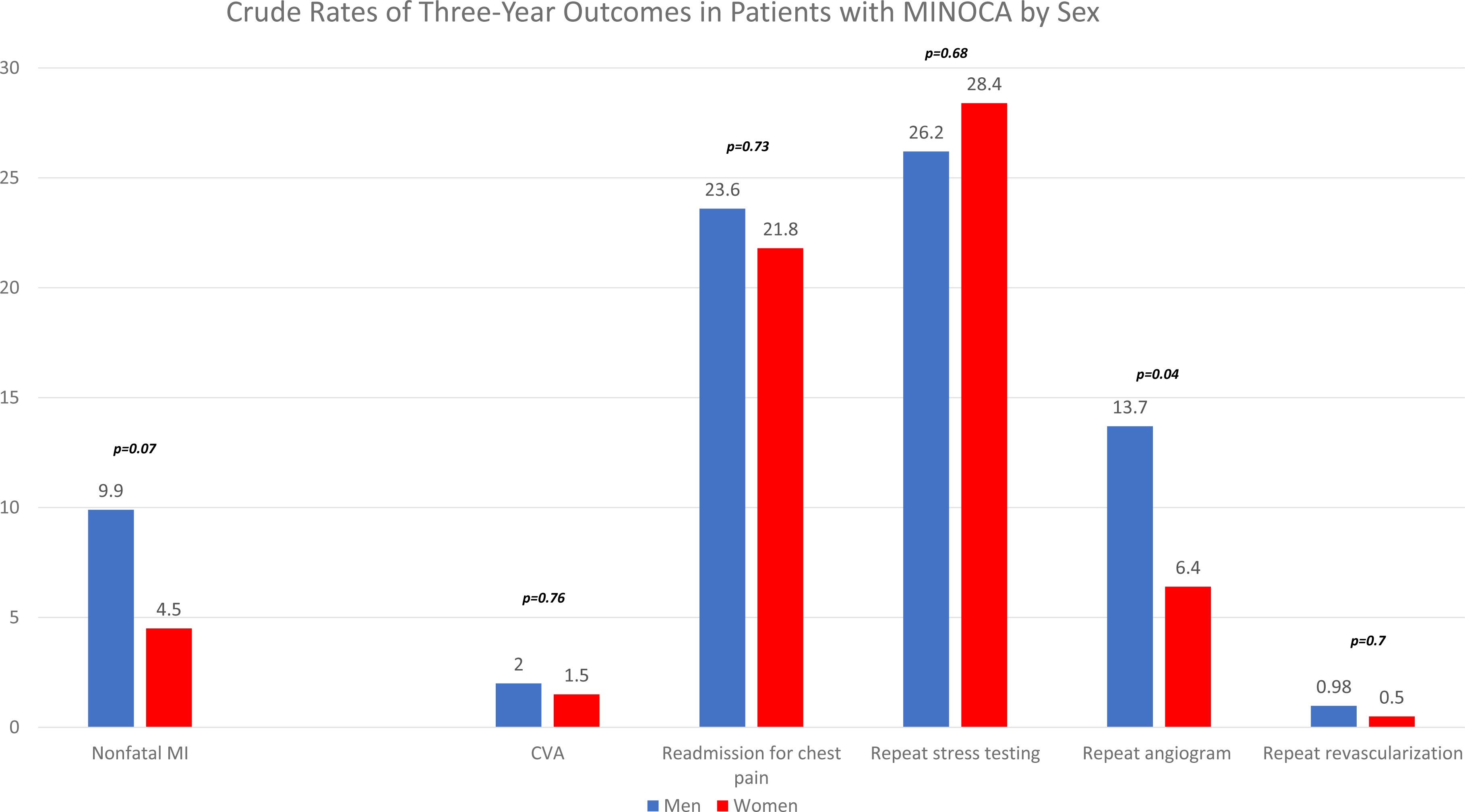
Crude rates of 3-year outcomes in patients with MINOCA by sex

**Figure 3.**
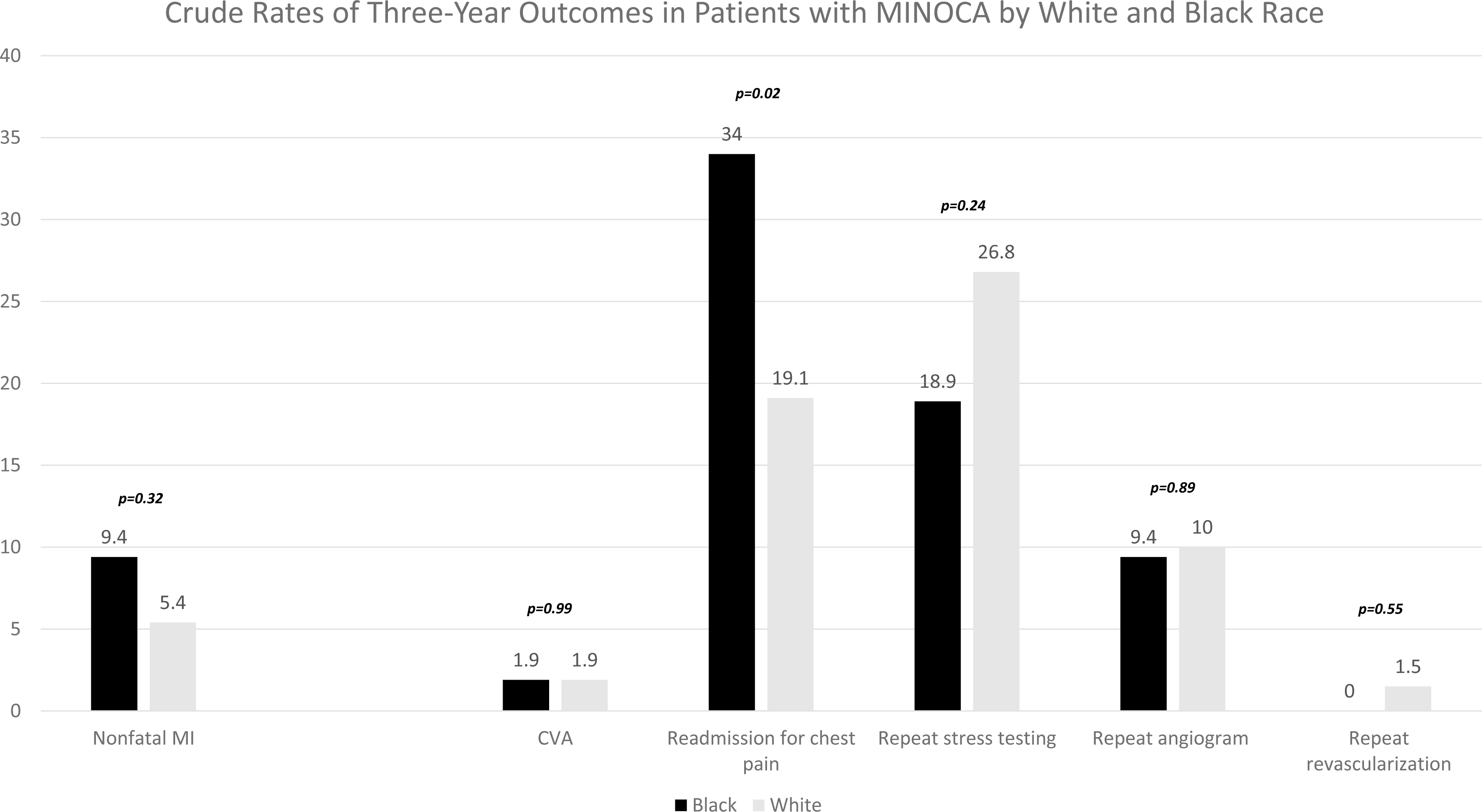
Crudge rates of 3-year outcomes in patients with MINOCA by race

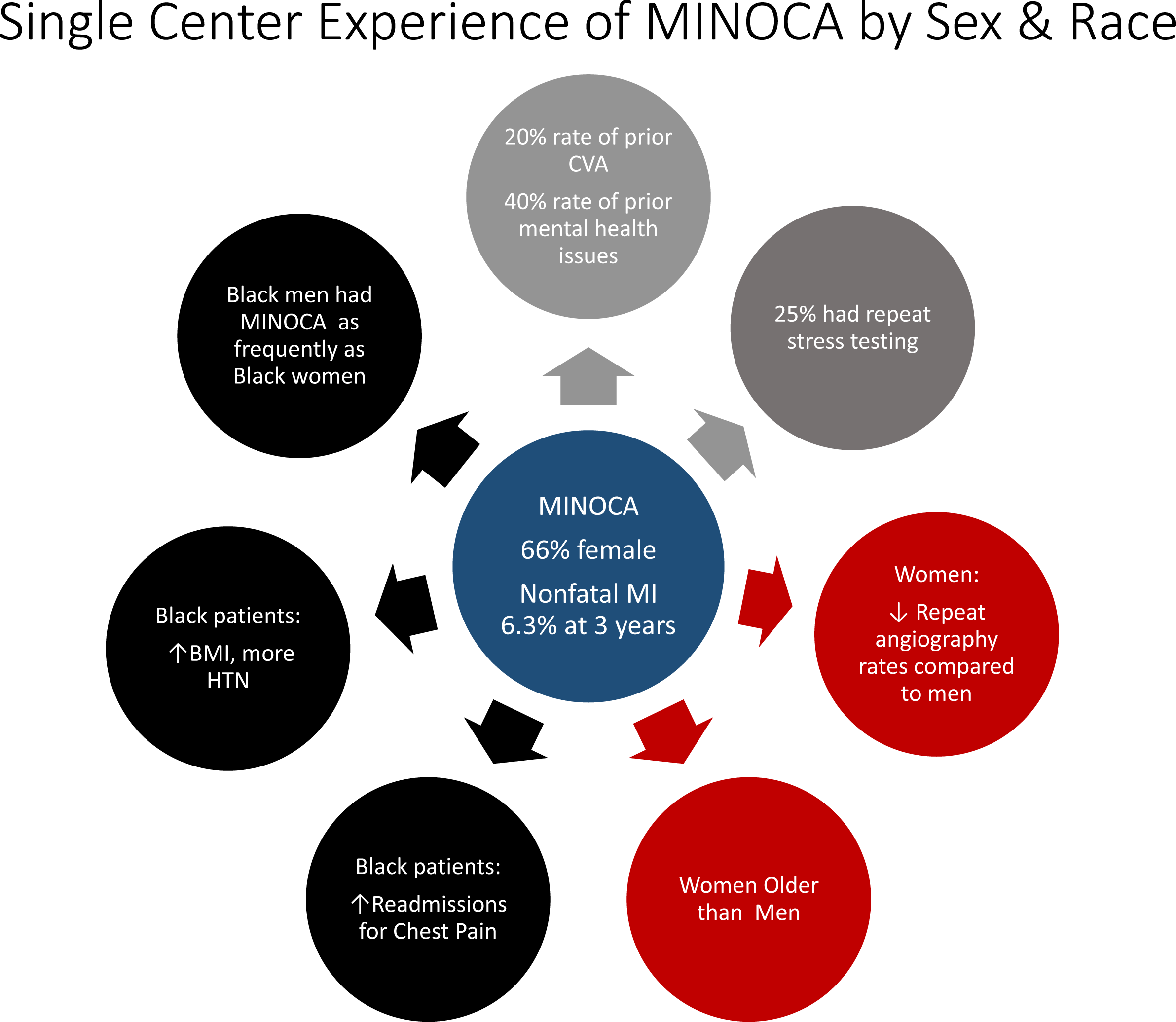

### Discharge Therapies

Almost three-quarters of patients (72.7%) were referred to cardiologists at discharge, with women more likely to be referred than men (72.7% vs. 64.7%, p=0.026) and no differences by race. Discharge medications showed that nearly half of patients were prescribed aspirin (49.3%), statins (44.1%), and beta-blockers (52.6%), with fewer receiving P2Y12 inhibitors (15.1%) or angiotensin-converting enzyme inhibitors (27.6%). Women were more likely than men to be discharged on beta-blockers (56.9% vs. 44.1%, p=0.03), without any other significant differences by sex or race. Only 7.5% of patients were referred to cardiac rehabilitation at discharge.

### Multivariate Outcome Predictors

There were no independent predictors of nonfatal MI (**Table 3**). Black race (OR 2.31 [95% CI (1.06-5.03)] was associated with an increased risk of readmission for angina, while female sex was associated with decreased odds of repeat angiography (OR 0.36 [95% CI (0.14-0.90)] and current smoking was associated with increased odds of repeat angiography (OR 4.07 [95% CI (1.02-16.29)] along with hyperlipidemia (OR 4.65 [95% CI (1.22-17.7)]. There were no other significant risk factors associated with long-term primary or secondary outcomes.

**Table 3.** Multivariate analysis of predictors of major adverse cardiac events, recurrent myocardial infarction, stroke in follow up, readmission for angina, repeat coronary angiography, and referral to cardiology follow-up

|  | **Nonfatal MI**<br>**Odds Ratio**<br>**(95% CI)** | **Readmission for Angina**<br>**Odds Ratio**<br>**(95% CI)** | **Repeat Coronary**<br>**Angiography**<br>**Odds Ratio (95% CI)** |
| --- | --- | --- | --- |
| **Female sex** | 0.46 (0.16-1.32) | 1.19 (0.61-2.34) | 0.36 (0.14-0.90) |
| **Black race** | 1.45 (0.41-5.11) | 2.31 (1.06-5.03) | 0.71 (0.20-2.51) |
| **BMI** | 0.99 (0.92-1.05) | 1.04 (1.00-1.08) | 0.98 (0.93-1.04) |
| **Age** | 0.98 (0.94-1.02) | 1.01 (0.98-1.03) | 0.96 (0.93-1.00) |
| **History of CVA** | 0.50 (0.03-9.07) | 1.06 (0.29-3.85) | 1.48 (0.28-7.84) |
| **Hypertension** | 0.49 (0.03-8.12) | 0.89 (0.25-3.15) | 0.59 (0.10-3.45) |
| **Hyperlipidemia** | 0.72 (0.06-9.23) | 1.03 (0.32-3.27) | 4.65 (1.22-17.7) |
| **Current smoker** | 2.98 (0.79-11.27) | 2.42 (0.89-6.59) | 4.07 (1.02-16.29) |
| **Previous smoker** | 0.92 (0.26-3.29) | 1.89 (0.92-3.89) | 2.40 (0.73-7.94) |
| **COPD** | 1.08 (0.08-14.25) | 0.83 (0.21-3.24) | 3.48 (0.87-13.94) |

**Abbreviations:** MACE, major adverse cardiac event; BMI, body mass index; CVA, cerebrovascular accident; CI, confidence interval; COPD, chronic obstructive pulmonary disease; MI, myocardial infarction;

## Discussion

This study identifies several important and novel findings about MINOCA. First, this study finds that men affected by MINOCA are less likely to be discharged on beta-blockers and referred to cardiology follow-up, and more likely to undergo repeat angiography than women with a numerically double the rate nonfatal MI in men compared to women but no difference by race at 3 years. Second, this study adds new information about race, as nearly half of Black patients with MINOCA were men, and Black race was associated with readmission for angina. Although prior studies have identified that women are more likely than men to present with MINOCA^1, 3^, without sex differences in mortality or MACE^4-6^, there has been very limited data reporting any racial differences in MINOCA^3^. We have demonstrated in this cohort that among Black patients, there was a nearly equal prevalence of MINOCA by sex. Third, there was a high prevalence of thromboembolic disease in this population, with almost 20% having a history of CVA and a 1.0% rate of CVA during the index hospitalization. Fourth, rates of use of cardiac magnetic resonance imaging in this cohort were low at 6.3%, as were rates of referral to cardiac rehabilitation at 7.5%. Finally, there was a high prevalence of mental health disorders in our population, with over 40% of patients diagnosed with anxiety, depression, or PTSD.

This study raises interesting questions regarding the long-term prognosis of patients with MINOCA. Only 6.3% had a repeat MI (largely repeat MINOCA episodes), but many patients had recurrent symptoms that led to additional hospital admissions and testing. Almost a quarter of patients were readmitted for chest pain or shortness of breath, over one-quarter of patients underwent repeat stress testing, and nearly 9% had repeat angiograms. Black patients were more likely to be readmitted, Black men were more likely to be referred for repeat angiogram, and all women were more likely to be medically managed. Despite these repeat ischemic evaluations and re-hospitalizations, rates of revascularization were very low at less than 1%. Taken together, these findings suggest inadequate symptomatic management of patients with MINOCA in long-term follow up despite low MACE rates. In prior research from the Women’s Ischemia Syndrome Evaluation (WISE), the estimated lifetime costs of non-obstructive coronary artery disease are 75% of the costs of obstructive CAD^7^. However, despite the costs associated with non-obstructive CAD, evidence-based therapies including cardiac rehabilitation, which have been shown to improve physical health scores and MACE in MINOCA patients^8^, are infrequently prescribed, as this study confirms.

Important finding in this cohort are the high rates of prior CVA at nearly 20%. The in-hospital rate of CVA at 1.0% was also unusually high and much higher than the expected rate of 0.05-0.1% with diagnostic coronary angiography^9^. In fact, it was higher than the <1% rate in patients presenting with ST elevation myocardial infarction^10^. One potential explanation for this could be an underlying predisposition to thrombophilia in patients with MINOCA. The mechanism of MINOCA remains unclear, but one pathophysiologic mechanism involves thrombus formation with subsequent lysis. This results in an apparently normal angiogram, but leads to a prothrombotic state predisposing patients to MINOCA^9^. This has been explored in several small studies, demonstrating thrombophilia disorders to be unusually high in MINOCA patients, ranging from 14-24%^11, 12^.

Another important finding in this cohort is the high prevalence of comorbid anxiety, depression, and PTSD despite excluding Takotsubo syndrome from this population. The association of mental health and cardiovascular disease has been described for over a century in medical literature^13^, with mental stressors shown to induce ischemia on myocardial perfusion imaging, despite many patients having no defect with conventional exercise or pharmacologic stress testing^14, 15^. The pathophysiology of mental stress ischemia appears different from that of conventional ischemia, with invasive studies showing signs of coronary endothelial dysfunction including decreased coronary blood flow and impaired vasodilatory response to acetylcholine during times of mental stress^16^. In prior MINOCA cohorts, there appears to be a high prevalence of comorbid psychologic disorders with estimates of up to 30%^17^. Anxiety and depression have also been noted to be independent risk factors for long-term mortality among Chinese patients with MINOCA, highlighting a possible connection between the pathologies^18, 19^. It is notable that the Black patients in this present urban cohort were actually less likely to have mental health disorders. Mental health disorders are commonly undiagnosed in Black patients, raising the possibility that the actual number of patients with psycho-emotional disorders is higher^20, 21^. Previous studies suggest a female predominance of MINOCA in White populations. In our cohort Black men are as likely as Black women to have suffered from this condition. Studies document a disproportionate burden of psychosocial stressors and allostatic load on Black men in the US^22^. Resultantly, Black men may be at higher risk for MINOCA and recurrent angina than White men, and a higher burden of psychosocial stressors may be causative. This should be assessed in larger, prospective studies.

Future studies raised by this analysis are the comparison between the incidence of mental health disease in patients with MINOCA versus obstructive CAD, evaluation of the association of CVA with MINOCA presentation, and assessment of the rates and long-term outcomes of patients referred for cardiac rehabilitation. Given the findings that the use of cardioprotective medications was low in this study at less than half of all patients, future studies should evaluate medical therapies including beta-blockers, statins, and antiplatelet agents on medium- and long-term outcomes.

### Limitations

This study is representative of a single urban academic medical center, which limits its generalizability. The initial medical care of MINOCA, the prescribed treatments, and follow-up patterns are likely representative of this single institution. Management upon discharge, outside of the hospital, was also not captured in this study. Additionally, patients may have died or had events in other health systems during follow-up that could not be included in our data, likely resulting in an underestimation of event rates. Although angiograms were reviewed to ensure MINOCA classification in these patients, these were not formally adjudicated by a core lab to specifically differentiate those patients with non-obstructive coronary artery disease from truly normal coronary arteries, which may effect outcomes by mechanism. Finally, the retrospective nature of this study raises the possibility of confounding and misclassification bias.

## Conclusion

This study adds novel information about sex, race, and underlying comorbidity risks for patients diagnosed with MINOCA. While MINOCA is classically described as a disease affecting women, this study confirms this pattern in White patients but identifies that Black men are equally as affected as Black women. There were high rates of recurrent hospitalization among Black patients and repeat ischemic testing among all patients, indicating high rates of symptom recurrence. Additionally, men are less likely to be discharged with beta-blockers and cardiology follow-up than women, suggesting that the diagnosis in men may be overlooked compared to women despite improving awareness around MINOCA management. Although racial differences were noted in the prevalence of CVA and mental health disorders, overall prevalence was quite high, raising the possibility of underlying thrombotic or neurohormonal mechanisms leading to MINOCA that should be evaluated in future studies.

## Data Availability

Data is Yale data accessed after IRB approval for retrospective review. No external datasets used.

## Author Contributions

GE: data extraction, data analysis, drafting of the manuscript, critical revision for important intellectual content

CH: data extraction, data analysis, drafting of the manuscript, critical revision for important intellectual content

TB: data analysis, drafting of the manuscript, critical revision for important intellectual content MK: data extraction, data analysis, drafting of the manuscript, critical revision for important intellectual content

MG: data analysis, critical revision for important intellectual content

QC: critical revision for important intellectual content, final approval of the manuscript submitted

HRR: critical revision for important intellectual content, final approval of the manuscript submitted

MG: critical revision for important intellectual content, final approval of the manuscript submitted

AV: critical revision for important intellectual content, final approval of the manuscript submitted

SEA: design of the protocol, data extraction, drafting of the manuscript, critical revision for important intellectual content, final approval of the manuscript submitted

